# Student and staff experiences of a novel curriculum-based peer support intervention (“study groups”) to support the mental health and wellbeing of postgraduate taught students

**DOI:** 10.1101/2024.02.23.24303245

**Authors:** Tayla McCloud, Tiffeny James, Sarah Rowe, Jonathan Huntley, Gemma Lewis, Claire Callender, Sonia Johnson, Jo Billings

**Author notes:** Corresponding Author Professor Jo Billings, Division of Psychiatry, University College London, Wing A, 6^th^ Floor, Maple House, 149 Tottenham Court Road, London, UK, W1T 7NF.

## Abstract

**Background:** In recent years there has been increasing concern for the wellbeing of higher education students, and institutions are under pressure to act. Loneliness and social isolation appear common among students, particularly postgraduate taught (PGT) students, and are linked to adverse outcomes such as depression and abandoning studies. We have in place a novel curriculum-based peer support intervention (“study groups”) that may help support the mental health and wellbeing of postgraduate taught students.

**Aim:** This study aimed to explore student and staff experiences of the study groups and their perceptions of how they may influence social cohesion, loneliness and wellbeing.

**Methods:** Qualitative semi-structured interviews were conducted with students and staff to explore their experiences and views of the study groups intervention. Transcripts of the interviews were analysed by the study authors following the principles of reflexive thematic analysis.

**Results:** We completed interviews with 20 students and five staff members. We found that students valued the study groups as a way to make friends, improve feelings of connectedness and cohesion, and receive emotional support. The shared experience of group members was key, although completing joint assignments could be a stressor for some.

**Conclusions:** This research suggests that this is a promising intervention to support the mental health and wellbeing of higher education students, and to reduce loneliness and improve social cohesion.

## Introduction

In recent years there has been increasing concern for the mental health and wellbeing of higher education students. In the UK, the prevalence of depression and anxiety in this population has been reported at 35% and 42% respectively, with five times as many students disclosing a mental health condition in 2015/16 than 10 years prior [1, 2] During this period, most higher education institutions reported increased demand for counselling services, with many struggling to keep up [2]. The possible consequences of poor mental health among students include impaired academic performance, abandoning studies, and suicide [3, 4]. Institutions are therefore under pressure to act to support and improve students’ mental health and wellbeing, focusing on prevention as well as intervention [5].

Socialising is a core aspect of university life and key to students’ wellbeing. However, many students experience loneliness and at university young people are separated from their usual support systems and social networks [6]. Increases in student numbers in the UK mean students may struggle to make friends among larger cohorts, and staff have less time to support students [7]. Loneliness is a strong predictor of poor mental health in students in quantitative data [8] and in qualitative interviews, social relationships are the most commonly reported stressors [9]. Experiencing loneliness at university is linked to adverse outcomes including depression, anxiety, self-harm and abandoning studies [10, 11]. Social support can improve wellbeing and protect against mental health problems, likely by enhancing students’ ability to cope with stressors and decreasing isolation [12]. This is therefore an important area of focus for institutions looking to tackle concerns around student mental health.

Among higher education students, postgraduate taught (PGT) students face a particular set of challenges, including more intense workloads and increased personal responsibility and independence [13, 14]. Shorter course lengths mean building relationships and social cohesion can be difficult for PGT students [15]. Qualitative research has highlighted that feeling a sense of belonging socially, building informal support systems, and having people to ask for information and advice are important for students’ wellbeing, but can be missing in PGT study [14, 16]. For students experiencing mental health problems, peer support may be more important than other sources of support [17]. This suggests that social interventions may be valuable in tackling loneliness and improving wellbeing among PGT students [8].

In the Division of Psychiatry at University College London (UCL), PGT students are placed in ‘study groups’ for academic, pastoral, social and organisational purposes. The study groups are used to practically manage approximately 120 students joining the programme through three interlinked MSc courses each academic year. It was also hoped that they would facilitate staff meeting students and students getting to know and learning from one another, with benefits for students’ wellbeing. In this way, these study groups can be considered a curriculum-based social intervention focused on peer support, fostering group memberships, and allowing students to connect with those facing similar challenges [8]. All students are placed into study groups. Groups comprise approximately 10-12 students, with a spread of international and UK students in each group. Part-time students are placed together and, in some cases, students taking similar modules are grouped together. Each group is assigned a study group lead who is an academic in the core teaching team, and a co-lead who is a PhD student or researcher in the Division and often a graduate of the Master’s programme. The leads and co-leads are intended to provide pastoral and academic support for the students. Students attend lectures and seminars with their study group in the first term, complete assignments together as a group, and meet regularly with study group leads and co-leads throughout the academic year. Study group meetings focus on topics such as academic tasks, wellbeing and career aspirations.

This study aimed to explore student and staff experiences of the study groups and their perceptions of how the groups may influence social cohesion, loneliness and wellbeing. This study took place during the COVID-19 pandemic and as such also provides some insight into students’ experiences of the study groups during remote learning.

## Methods

### Design

Qualitative semi-structured interviews were conducted with MSc students and staff to explore their experiences and views of the study groups intervention. All procedures were approved by the UCL Research Ethics Committee (ref: 8227/004).

### Participants

Students’ experiences were the focus of the study, with staff perspectives used to complement these. Students were eligible to take part if they had been enrolled on one of the three MSc courses on the UCL Division of Psychiatry Master’s programme in the 2019-20 or 2020-21 academic years. Staff were eligible if they had been a lead or co-lead for the study groups for both academic years.

We used purposive sampling to achieve a diverse range of participant characteristics and experiences. Students were recruited via an invitation email sent to those studying on the MSc programme at the time (‘current’ cohort) and posts by JB on Facebook groups for the current (2020/21) and previous (2019/20) cohorts. The COVID-19 pandemic lockdowns and online learning were in place from approximately halfway through the first cohort’s programme, and from the beginning of the second cohort’s programme. Staff were recruited via an email to all leads and co-leads who had been involved in the study groups for both academic years, excluding the study authors. Potential participants were invited to contact the lead researcher (TM) via email. Volunteers were sent the Participant Information Sheet and asked to complete some socio-demographic questions. All participants gave informed consent by email before taking part.

### Data collection

We conducted semi-structured interviews using two interview schedules: one for students and one for staff. Both schedules included similar questions about participants’ experiences of the study groups and views on how they may impact students’ wellbeing, social cohesion and loneliness (see supplementary information).

Before the interviews, all potential participants were asked their gender, ethnicity, and which study group they were a member of. Students were additionally asked their fee status (overseas/EU/home student), enrolment status (full-time, part-time or flexible/modular), and MSc course (dementia, research or clinical).

Interviews took place via Microsoft Teams online video call or telephone, depending on participants’ preferences. Interviews were conducted by TM or TJ, supervised by JB. Students from the 2019/20 cohort were recruited first and interviewed between November 12^th^ and December 18^th^ 2020. Students from the 2020/21 cohort were interviewed between February 5^th^ and March 1^st^ 2021.

Before starting each interview, participants were reminded that anything said was anonymous, and were encouraged to speak openly and honestly regardless of whether their experiences were positive or negative. Interviewers were supported by JB, a consultant clinical psychologist and experienced qualitative researcher.

### Data analysis

All interviews were audio recorded then transcribed verbatim by TM or TJ. All potentially identifying information was removed to protect participants’ anonymity. Participant numbers are used with illustrative quotes in this paper.

Data analysis was conducted by TM and TJ, in collaboration with and overseen by JB. All analyses followed the principles of reflexive thematic analysis. This involves identifying patterns and themes in the data, and requires researchers to consider their own context and how it influences data generation and analyses [18]. We coded data inductively, meaning the coding structure was derived from the data and not pre-determined by existing theories or research. Student and staff interviews were analysed simultaneously.

TM, TJ and JB read three different transcripts each, coding freely, then together discussed potential codes and possible themes. All transcripts were then imported into NVivo 12 [19] and coded by TM and TJ into the provisional coding structure. This was further developed and refined iteratively throughout the rest of the coding process and discussed regularly in study meetings between TM, TJ and JB. A final set of themes was developed from the coded data, revised with feedback from the wider study team and student consultation group, and finalised upon write-up.

### Coproduction

The study team is made up of mental health researchers and clinicians at various career stages. All are based within the UCL Division of Psychiatry and involved with the teaching and/or organisation of the Master’s programme and study groups. TM and TJ are graduates of the Master’s programme and had experienced the study group intervention themselves, as students and as co-leads. We reflect on the potential implications of this in the Discussion section. Neither TM nor TJ interviewed any participants who had been part of their study group.

We recruited a student consultation group to coproduce the research. This comprised four students from the 2019/20 Master’s programme cohort and one student from a UCL Master’s programme without study groups. The consultation group collaborated with the study team to draft the advertisement text and interview schedules, develop the coding structure, review developing themes, and review the present paper. The consultation group did not take part as participants.

## Results

### Participant Characteristics

Eight students from the 2020/21 academic cohort, twelve students from the 2019-20 academic cohort, and five staff members (two leads and three co-leads) were interviewed. Students were from twelve different study groups. All interviews were conducted remotely via online video call and lasted between 27 and 66 minutes. Participants’ demographic information can be found in Table 1. Broad categories are used for anonymity.

| **Sample characteristics (n = 25)** |  |
| --- | --- |
|  | N (%) |
| **Role** |  |
| 2019/20 student | 12 (48%) |
| 2020/21 student | 8 (32%) |
| Lead/co-lead | 5 (20%) |
| **Gender** |  |
| Female | 21 (84%) |
| Male | 4 (16%) |
| Other/prefer not to say | 0 |
| **Ethnicity** |  |
| White British | 8 (32%) |
| White Irish/White Other | 7 (28%) |
| Asian/Asian British | 7 (28%) |
| Mixed/Other | 3 (12%) |
| **Student-only characteristics (n = 20)** |  |
|  | N (%) |
| **Enrolment status** |  |
| Full-time | 20 (100%) |
| Part-time or flexible/modular | 0 |
| **Fee status** |  |
| Home | 8 (40%) |
| EU | 7 (35%) |
| International | 5 (25%) |

Of the 25 participants, 21 (84%) identified as female and eight (32%) were of White British ethnicity. Of the 20 students, 100% were enrolled full-time and eight (40%) were classed as home students.

### Overview of themes

Five main themes were derived from the data: ready-made go-to group, diversity of experience, investing in the group, connectedness and cohesion, and shared experience. These themes and the sub-themes are shown in Table 2. Overall, staff and students viewed the study groups intervention positively, with most students reporting a beneficial impact on their Master’s experience and wellbeing. Nevertheless, various factors shaped students’ experiences of these groups and the mechanisms by which they impacted on wellbeing, loneliness, and social cohesion.

**Table 2.** Themes and Sub-Themes.

| Theme | Sub-Themes |
| --- | --- |
| 1. Ready-made go-to group | Mandatory meet-ups<br>Familiar faces and meeting people<br>Point of contact |
| 2. Diversity within groups | Career stages and previous work experience<br>Culture |
| 3. Investing in the group – “get out what you put in” | Approach to work<br>Division of labour<br>Roles in the group |
| 4. Connectedness and cohesion | Luck of the draw<br>Keeping in touch<br>In-group and out-group<br>Lasting friendships |
| 5. Shared experience | Friendship vs work<br>Emotional support<br>‘In the same boat’<br>Joint learning |

#### 1. Ready-made go-to group

This theme describes how students’ wellbeing and loneliness was positively impacted by allocating students to smaller subsets of the larger cohort and giving them compulsory activities to complete together.

##### Mandatory meet-ups

Most participants thought it was helpful for the course to provide compulsory, regular, structured opportunities to work together and socialise, particularly at the beginning of the academic year:

> *“It felt so natural, like we were put into these groups and the relationships were formed so easily, they just sort of fell into place so quickly and I don’t know whether that would’ve been the case if we didn’t have the study groups… […] I think it really enhances the social integration on the course.”*
>
> *[Ppt 4, female home student, 2019/20]*

Mandated study group interactions were perceived as beneficial for ensuring there was always some interaction between students. Those who were lonely or less integrated still had to talk to other students. The general consensus was that the level of peer support and joint learning experienced would not have occurred naturally without organisation by the course leaders.

##### Familiar faces and meeting people

All participants cited how study groups helped to reduce loneliness and improve social cohesion by facilitating familiarity with peers and staff from the beginning of the course. This was described as beneficial for students’ wellbeing, particularly on a large Master’s programme that might otherwise feel overwhelming socially. One student told us:

> *“It’s much nicer to walk into a room knowing it’s full of people you know and you’re friends with and you’re all learning together, rather than coming in and they’re kind of strangers […] [That] has been really positive on wellbeing and it really helped lift moods… especially in a stressful environment.”*
>
> *[Ppt 4, female home student, 2019/20]*

The study groups created a “sense of belonging” and provided students with what one staff member called a “mini family” within the cohort. The groups enabled students to get to know staff on “first-name terms”, fostering a personal relationship with the Master’s programme. This helped students feel part of the community within the department and the wider University, which they may not have otherwise. One staff member told us:

> *“Coming to the division for a tutorial or for a study group meeting I think is a part of helping them feel like this is your place and this is the smaller thing within UCL that you belong to.”*
>
> *[Ppt 17, staff]*

The study groups were recognised by students and staff as an important source of social support in the context of the COVID-19 pandemic. Students described it as “almost impossible” to make initial contact with other students away from the study groups as a result of remote learning.

##### Point of contact

The study groups provided a designated point of contact where students could ask questions and receive information and advice from their peers. This was described as practically useful and helpful in alleviating anxiety:

> *“It’s always good to have someone to ask a question about a deadline or submission or anything like that. I think it kind of made the whole stress from a really intense Master’s course less stressful, because you felt like you weren’t in it alone.”*
>
> *[Ppt 11, female EU student, 2019/20]*

Study group co-leads, many of whom are programme alumni themselves, were described as an important and valued source of support. Co-leads helped alleviate anxiety and uncertainty by answering questions and signposting to information and support, which students noted was reassuring and helpful. Co-leads could also act as a “middle person” between students and the teaching team, and as an important conduit for feeding information about student wellbeing back to the University.

> *“If a student doesn’t feel good or feels anxious…having study group meetings with the co-lead and lead can be a chance for us as students to be heard and to make sure that the University knows what we are going through and what are our difficulties.”*
>
> *[Ppt 14, female EU student, 2020/21]*

#### 2. Diversity within groups

This theme describes how the study groups often bring together a diverse group of individuals with different experiences and backgrounds, which can be useful for learning and group cohesion.

##### Career stage and previous work experience

Diversity in study group members’ career stages and prior work experience was highlighted by several students. For some, this made the group was more engaging and helpful, as there was room for skill sharing around jobs and course content. However, differences in priorities, particularly between full time students and those studying part time alongside work, could create challenges in scheduling.

Often more work experience tallied with study group members being older in age. Several students told us that this affected the group dynamics:

> *“I think the fact that there were some older, more mature students helped in the sense that…having work experience or being a few years older sort of made those people maybe more nurturing of the younger ones… [But] even though I was the oldest in the group, I was also the one who felt the most out of my element because I hadn’t done psychology in so long, and so I also felt quite reliant on [the younger ones] in terms of reassurance seeking.”*
>
> *[Ppt 3, female home student, 2019/20]*

##### Culture

Having a mixture of home and international students within the study groups seemingly impacted integration and was good for personal development and cohesion:

> *“I’d say cohesion in a sense comes from diversity, of backgrounds and of different experiences. Because when you think of cohesion… you might think that [all being home students] would lead to more cohesion but I think having a group of different experiences can actually help to increase the chances that people in the group will get along well with each other.”*
>
> *[Ppt 3, female home student, 2019/20]*

However, study groups with more home students could be isolating for international students, and some sought friendships or support outside of their study group with those they could identify with better. Some students observed international students being quieter and less confident than home students in group meetings or when working on group assignments. There was a sense that international students may find the group challenging or stressful, especially at the beginning of the course. One international student described how the study groups could both reduce and highlight loneliness:

> *“In some ways the study group made me less lonely because it pushed me out from my comfort zone and meant that I needed to talk to people who are not from the same background as me. And after attending [group meetings] I would feel better and less lonely. But sometimes the study groups reminded me about loneliness because they are not from the same culture as me, and […] I felt a bit excluded because of the language issue”*
>
> *[Ppt 6, female overseas student, 2019/20]*

Similarities and differences between students’ previous courses and institutions impacted the groups. For example, some students told us that those who hadn’t studied in the UK before struggled with things like an unfamiliar grading system, which could increase feelings of loneliness, but having others in their study group with whom they could discuss this helped them bond.

#### 3. Investing in the group – “getting out what you put in”

This theme explores how members of the study groups came together to work as a team. The amount and way in which individuals contributed to the group and group assignments impacted the group’s cohesion and individuals’ wellbeing, positively and negatively.

##### Approach to work

Group assignments mostly did not contribute to students’ final grade, but required students to reconcile different personalities and approaches to work to produce something together. This was commonly discussed as a source of stress and anxiety, particularly at the beginning when students were also trying to make friends and avoid conflict. Students described feeling very aware of the “group atmosphere” that emerged out of this, which in itself could cause anxiety:

> *“At times, the study group can feed into that anxiety a little bit because my study group were very academic… sometimes if everyone else around you is too stressed, it can make me more stressed.”*
>
> *[Ppt 5, female EU student, 2019/20]*

It was sometimes difficult for students to reconcile their individual approaches to assignments and agree on the standard of work to aim for. This was expressed in terms of “trust” and “reliance” on others to ensure the work was completed to a good standard. Some students expressed difficulty accepting that group assignments might differ in quality from their individual work, and anxiety that this could hinder their own learning. Where students were aligned, cohesion improved, but where they were not, this could increase tension within the group and cause stress for individuals on both sides:

> *“That could cause a bit of conflict and tension, because people disagreed on the best way to go about stuff and [had] contrasting personalities”*
>
> *[Ppt 12, female home student, 2019/20]*

This generally improved as the year went on and more assignments were completed. Leads and co-leads could also be helpful, for example in tempering an intense atmosphere:

> *“[They were] very calm and level-headed and answered all our questions and I think that really helped because I’d say that my study group was a rather high energy bunch (laughing)…like ‘are we doing this right?’ and [co lead] would… reassure us that everything was okay. [They] really balanced out a lot of the more intense personalities in the group.”*
>
> *[Ppt 3, female home student, 2019/20]*

##### Division of labour

The most common challenge associated with study groups seemed to be around practically dividing up group work tasks, and several students were apprehensive about this from the beginning. Many students described the division of labour as unbalanced, often due to different levels of commitment and motivation within the group:

> *“I would say [the study group] had a very minimal effect [on my wellbeing], if anything, slightly negative, causing me stress. It felt like almost unnecessary stress for the [assignments] that didn’t count, because I was kind of like, ‘this doesn’t matter and I’m doing all the work for it, and everyone else just gets away freely with doing nothing’.”*
>
> *[Ppt 2, female home student, 2019/20]*

While students who did less work or easier parts avoided the stress of the increased pressure, some worried that they may have learned less from the assignments. Some groups worked well as a team:

> *“I was really surprised because everyone was working really hard and trying to do their part the best they could […] Usually with group work there’s always one person who does more and another person who doesn’t do anything but I didn’t feel like that was my study group at all.”*
>
> *[Ppt 11, female EU student, 2019/20]*

Interviews highlighted a lack of clarification of the role of the study group leads and co-leads among both students and staff. Some students suggested that study co-leads could have a positive influence on wellbeing if they encouraged teamwork and a fair division of labour, or problem-solved around this issue.

##### Roles in the group

The roles individuals took on in the group was a source of stress for many students. It could be challenging, especially in larger groups, if no one stepped up to take the role of leader, but equally, too many leaders could create a difficult working environment. In both cases, groups could be less cohesive, which could lead to anxiety and difficulties completing work. Some who did take the lead said they found it stressful to organise others and potentially take on more work. However, students who gravitated towards leadership often bonded with other like-minded peers. Having more outgoing “leaders” in a group was perceived as encouraging the group to be more social. Eventually, most groups found a way to slot into roles that worked well:

> *“No one really wanted to take the lead per se, and everyone had quite busy schedules so I think it was a bit stressful to try and find a time that we could all meet and assign things, and no one really wanted to step up to that leader role because… I feel like once you step up to it you kinda have to maintain it… It did work out well in the end but it was quite stressful.”*
>
> *[Ppt 9, female home student, 2019/20]*

#### 4. Connectedness and cohesion

Study groups provided an opportunity for students to connect with peers, and the quality of those connections impacted students’ wellbeing and loneliness. Connectedness and cohesion varied across groups and was influenced by several factors.

##### Luck of the draw

Due to their random allocation, the success of each study group was perceived to a large degree as dependent on luck. Students described apprehension about this, knowing that there is not much that can be done if a group doesn’t “click”. Being in a group that worked well together, got along socially, and was cohesive was beneficial for students’ wellbeing and loneliness. Students in groups that got along well described feeling “fortunate” and “lucky”.

However, a group that lacked these attributes and the related benefits could be detrimental for students’ wellbeing, with awkward or tense interactions during group work. One student who experienced two different study groups told us:

> *“It is sort of a luck of the draw who you’re put with. I was quite stressed out in term one, having to gather everyone together and it was only one or two of us who were doing that, and that I would say was a source of stress and a bit of anxiety for me […] Whereas my second group it was a completely different dynamic and I think in that sense, my wellbeing has improved and stress levels have decreased.*
>
> *[Ppt 15, female home student, 2020/21]*

This concept also applied to the allocation of leads and co-leads, with students aware that groups received varying amounts of involvement.

##### Keeping in touch

This sub-theme explores how students kept in touch with one another as the mandatory group tasks lessened throughout the academic year. If students managed to keep in touch when it was no longer compulsory, the group could form a closer bond and become more of a “social space”, helping to facilitate greater connection between students. Several students said that they appreciated having a co-lead who made the effort to keep in touch with them, as they felt more supported and connected to the course.

Students from the 2019/20 academic year transitioned to remote learning halfway through the course, so keeping in touch became even more important and more challenging. The risk of feeling lonely in this context was mitigated for some by maintaining regular contact with their study group. Students found this “reassuring” as it provided an opportunity for peer support and helped them stay connected to the course.

Many students said that setting up a Whatsapp group together was key to staying connected. However, as online socialising became more important, it could be difficult to keep in touch with those who didn’t engage in this. Several students noted those who were already less involved in the group were more likely to remain isolated as there were fewer opportunities for casual social interactions and direct communication:

> *“I still think it’s very different compared to sitting in a lecture with your coursemates and then going to grab coffee.”*
>
> *[Ppt 1, male overseas student, 2019/20]*

2020/21 students started the course during COVID-19, so while the study groups’ social side was helpful to an extent, trying to initiate, develop and maintain friendships entirely online was difficult:

> *“Because it was all via Zoom it wasn’t like people would stay after and have a bit of a chat […]. I think other people did feel similarly in the sense that everyone did feel lonely and a bit isolated. But would you really want to sit on a Zoom call for another half an hour? It was just that whole predicament of people wanting to make friends but didn’t know how.”*
>
> *[Ppt 22, female overseas student, 2020/21]*

##### In-group and out-group

There was a sense among students that, as study groups dictated much of their time and social interactions, it could be difficult to socialise with other members of the cohort outside this. There seemed to be a feeling that people were obliged to be friends with their study group and were otherwise “missing out” on this aspect of the Master’s programme. One student told us:

> *“Personally, I don’t feel like [my group] helped me to integrate, I actually think it made me feel more left out because you can see other study groups that are bonding a lot […] and I’m like ‘people from my study group just barely speak’ […] and then I kind of felt like I couldn’t interact with people from other study groups because they were already bonded so well.”*
>
> *[Ppt 11, female EU student, 2019/20]*

Groups that had bonded well together were described as a “bubble” that was difficult for those outside to penetrate. Most students said people didn’t tend to socialise outside their study groups. In groups that did bond together well, students felt that a key benefit was having a subset of the larger Master’s programme to connect with. One student explained how the atmosphere of their study group wouldn’t otherwise be experienced:

> *“I ended up in a really good, supportive study group who I’m still in touch with now, and I never really felt like I was in competition with anyone. I think as it is quite a competitive Master’s and I did feel that from other people outside of my study group, I was quite grateful to have this sort of oasis of non-competition.”*
>
> *[Ppt 3, female home student, 2019/20]*

##### Lasting friendships

Study groups facilitated the forming of friendships that lasted beyond the course, with social meet-ups and valuable career-related support ongoing:

> *“We maintained contact throughout the whole year and still now, we’ve been meeting up and going out for lunches and dinners […] It was quite useful with speaking to other people about what sort of jobs they’re applying for and we had a go at reading over each other’s applications for jobs and making suggestions for one another and supporting each other on the next part of the journey after the MSc. […] I can’t see us losing touch because they’ve been really good friends.”*
>
> *[Ppt 4, female home student, 2019/20]*

This was generally described as unexpected; students told us socialising isn’t usually the focus of a one-year Master’s and likely wouldn’t have happened without the study groups. Study groups opened students up to the possibility of making lasting friendships, rather than just being there to learn, which supported wellbeing and made the course more enjoyable. Some students explained that friendships were partly born out of the amount of time study groups are required to spend together, and partly due to experiencing an academically demanding year together. However, watching close friendships develop in the group could make it more difficult for members who were not part of that.

#### 5. Shared experience

This theme explores how the shared experience of working on group assignments and completing a Master’s together impacted students’ wellbeing. Time spent together could be a source of enjoyment and comfort for students bonding over shared challenges, or could be a further stressor.

##### Friendship vs work

Interviews explored the balance between friendship and work within study groups, and the extent to which groups had an academic or social focus. This differed between groups and was influenced by the personalities of group members. This balance (or imbalance) could affect whether the study group impacted individuals’ wellbeing and loneliness, with groups that got along well and socialised together having a more positive impact.

This fluctuated throughout the academic year. With more prescribed time spent together in the first term, students had more opportunity to build bonds, but were also busier with assignments. Some students were unclear about the purpose of the study groups, and for some, friendships were viewed as accidental by-products of the situation rather than actively intended by the course organisers:

> *“As time went on I realised that I was getting on really well with people in the study group and that I [felt] like I could reach out to them about other things as well and it wasn’t just academic. So that probably shifted quite quickly, but I think for the first meeting…I don’t think I understood it as a social space, or that it was going to be for wellbeing purposes or anything like that.”*
>
> *[Ppt 7, female EU student, 2019/20]*

This concept also applied to meetings with study group leads and co-leads, and the extent to which they focused on professional or personal topics, which differed widely between groups. Some students felt that meetings with only the co-lead were more beneficial as they could create a more friendly, informal space leading to more group cohesion. Meetings with senior leads were often seen as more formal and work-focused, so students were less comfortable discussing their wellbeing. One lead told us:

> *“I see my role as somebody they can connect with as a member of the MSc team, and also academic guidance, a bit on the pastoral stuff…but I think actually the co-leads have a more important pastoral role […]. I can sort of talk to them and encourage them to open up and share their problems and so on but I think there is a bit of a barrier.”*
>
> *[Ppt 17, staff]*

##### Emotional support

Study groups were seen as a valuable source of emotional peer support for students, on a group and individual level as well as with leads and co-leads. As well as the COVID-19 pandemic, at different times various students struggled with their workload, found assignments stressful, or dealt with mental health problems. The study groups were seen as a constant that could be relied on throughout:

> *“We had a group chat and we would use it for different kinds of support. If people were really stressed about an assignment, we could use the group chat to manage people’s stress and… we also touched upon personal things. […] There were so many different events […] and our study group was always there, I could get support from them if I ever needed.”*
>
> *[Ppt 1, male overseas student, 2019/20]*

Several students experienced mental health problems during the course or had supported someone who had, and many highlighted that study groups provided space to discuss this. Students in cohesive study groups checked on one another, and if someone was struggling or less engaged than usual it would be picked up on by their peers. Students also sought support from one another in relation to academic work, but only in groups that had formed friendships:

> *“I sought a lot of comfort and reassurance from my study group […] When I had to present for journal club, I was really terrified, and one of the [people] in the study group volunteered to do it at the same time as me…[they] saw how nervous I was, so [said] ‘don’t worry, I’ll be there with you’, so I really appreciated that support.”*
>
> *[Ppt 3, female home student, 2019/20]*

This also applied to relationships with staff, with many students wanting more emotional support from their study group leads and co-leads. Students reported a cycle whereby less involvement from staff created less opportunities to build rapport and therefore less personal and emotional disclosure. This then created a more formal atmosphere whereby students felt they could only discuss academic or course-related issues with staff. This was a common sentiment regarding leads, often in contrast with co-leads, but was sometimes true for co-leads as well.

##### ‘In the same boat’

Students frequently described the study groups as a place to validate and bond over shared experiences. There was a sense of “camaraderie” and “solidarity” among the group when studying together and discussing academic work, particularly when the workload was challenging. Students were facing similar difficulties and could encourage one another and empathise with their current situation in a way that their friends and family couldn’t, which helped people feel less alone:

> *“It’s not necessarily about them being your friendship group, but [the study group] gives you a valuable connection to people that are doing the same assignments as you, having the same stresses as you. I definitely felt that when I had to miss out on things… there was that sense of, well there’s also some other people who are working on weekends or stressed about an assignment (laughing), I’m not the only one in the world who’s locked into library mode.”*
>
> *[Ppt 10, female home student, 2019/20]*

This was especially true during the COVID-19 pandemic, where study groups provided “*some tangible solidarity beyond the imageless, video-less people on Zoom*” [Ppt 19, male EU student, 2020/21]. Another student said:

> *“I found it really hard, and having other people that I could text and go ’I feel like I’m struggling, how are you getting on with everything?’ and realising that actually they feel the same, it was really helpful. […] If I didn’t have that, I think it could’ve been really detrimental to both my learning and my wellbeing.”*
>
> *[Ppt 4, female home student, 2019/20]*

The fact that the co-leads were also past students was described as helpful for being able to relate to them and feel validated by them. Co-leads provided an example of someone successfully completing the Master’s, which was reassuring to students when the course was challenging.

##### Joint learning

Students told us about the benefits of learning from and alongside one another in group assignments both for their own individual learning and skill development, as well as for their wellbeing. When groups worked well together on formative tasks, this built cohesion and allowed students to learn from one another without the pressure of summative assignments:

> *“You’re not left fully on your own […] everyone’s come from different universities so some people may have not done something before and that can be really scary… so the fact that you can bounce ideas off each other and get feedback, that was so helpful.”*
>
> *[Ppt 5, female EU student, 2019/20]*

Group work also introduced an enjoyable social element to what could have otherwise been a solitary learning experience. Study group tasks were described as a welcome change from lectures for this reason, especially when learning remotely:

> *“Journal club, it was just brilliant, because it was four people, we could all bring our personalities to the table, we were making little remarks and that’s the aspect that I miss so much. It was all on zoom, we never met, but it still felt like ‘I’ve made some friends and here’s something I like and people I can talk to’ and that was really nice.”*
>
> *[Ppt 22, female overseas student, 2020/21]*

However, for groups that weren’t functioning well, the group assignments could hinder individuals’ learning, cause social tension, and negatively influence wellbeing. For example, rushing to complete tasks last-minute without oversight of the whole assignment could cause added confusion and stress.

## Discussion

In this study, we sought to explore student and staff experiences of a novel study groups intervention, with a focus on social cohesion, loneliness and wellbeing. We found that students valued the study groups as a source of connection, camaraderie, practical information and learning, and a link to the staff and wider department. They generally saw them as beneficial to their wellbeing. The study groups were described as generating a sense of belonging and a space for community building and informal support, all of which have previously been established as important for both learning and wellbeing [14,16,20]. Study groups connected students who were facing similar demands academically (and perhaps emotionally), which has previously been noted as an active component of peer support in higher education [8]. Many students formed friendships and supported one another in ways that they felt they would not have without a formal structure in place, perhaps due to the transient nature of PGT study [16]. This is also likely to be beneficial for students’ learning as well as wellbeing [21]. Nevertheless, for some people, the study groups also came with stressors such as navigating group dynamics and dividing group work. Completing academic tasks together created spaces for socialising and peer support. However, mandatory time together could be challenging if students did not get on well, and group assignments could be a source of tension. This could be due to differences in expectations and prior experiences of higher education [13].

There is an element of chance that meant that some groups were less cohesive - either socially, when studying, or both. Implementing study groups as a mandatory part of the MSc course meant that students saw them as an academic responsibility, and the social and wellbeing aspects often grew organically. This has been noted as helpful in maximising engagement with peer support [22] but it meant that some groups remained academic in focus. For students in these groups, the intervention could be a barrier to making friends, as there was less socialising outside of the study groups.

### Strengths and limitations

We deliberately sought to recruit a diverse range of experiences and perspectives for this study. We spoke to students from two cohorts as well as study group leads and co-leads, and the gender balance is in line with the student body. We were, however, unable to recruit any part-time students, perhaps because many balance employment and family life on top of their studies. Our findings therefore do not reflect the experiences of part-time students who may experience the study groups differently, for example as an added responsibility and potential source of stress.

All members of the study team had involvement with the study groups, and the two researchers who conducted interviews and data analysis had experienced the intervention as both students and co-leads. Whilst TM & TJ did not know participants directly, both may be known to students as MSc programme staff, which could mean they felt hesitant to discuss negative experiences of the study groups. We perceived, however, that being current PhD students and previous Master’s programme students improved rapport with and personal disclosure from participants, as the interviewers could understand and empathise with situations they were describing. While lived experience gave us an in-depth insider perspective during data analysis, we may have blind spots due to our familiarity with the topic and potential difficulty being impartial. Our consultation group provided a valuable additional validity check when they agreed that our findings reflected their experiences.

All students were studying for an MSc programme on mental health. It may be that students were particularly aware of one another’s wellbeing and in a position to provide emotional support to one another due to the nature of the course content and students’ knowledge of and interest in mental health. It is uncertain how transferable our findings are outside of this MSc programme – for example, for other PGT students, undergraduates, PhD students, smaller cohorts and longer courses.

Data collection for this study took place when remote learning was in place owing to the COVID-19 pandemic. All students interviewed had experienced significant disruption to their studies. This likely shaped their social and pastoral experience of the Master’s programme and the study groups intervention. We believe that many benefits of the study groups were heightened during this time - for example, providing a link to staff members and a forum for emotional support and friendship. Nevertheless, our conclusions may have differed had we interviewed students in a more typical year.

### Implications

Whilst there is nothing to suggest that our findings would not be applicable to alternative settings, future research could build on our findings by evaluating study groups on different types of courses – for example, smaller MSc cohorts, different disciplines, and undergraduate programmes. To complement the present study, it would be beneficial to quantitatively evaluate the effectiveness of the intervention at improving students’ mental health and wellbeing by, for example, comparing students on two otherwise similar courses with only one cohort receiving the intervention.

We have provided details of and findings related to our implementation of this intervention. For example, we have highlighted the role of leads and co-leads in helping students manage group assignments, such as by setting expectations around equitability, the standard of work and independence. Students confirmed positive aspects of the intervention that may be key to their success, such as bringing groups together from the very first day and mandating meet-ups. The need for details on delivery methods of peer support interventions has previously been noted [23].

Evidence to support curriculum-embedded interventions targeting student wellbeing is limited and curriculum-embedded peer support interventions are rare [24]. We have shown that study groups are a promising, novel, curriculum-embedded peer support mechanism. Our findings suggest that there are potential benefits for improving students’ mental health and wellbeing, reducing feelings of loneliness, and improving social cohesion within the cohort. We have shown that the groups are acceptable to students, and may also be beneficial for students’ learning. Similarly, this intervention is valuable in being focused on prevention [25]. Study groups are cost-effective to implement, with a minimal time cost and low effort for course organisers for potentially large benefits. As a result, this intervention is likely to be of benefit to other course organisers and higher education institutions [7].

## Data Availability

Due to the personal and in-depth qualitative accounts given by participants in this study, and in line with our ethical approval, data has not been made publicly available.

## Supplementary Information

S1 Interview guide for students

S2 Interview guide for staff

## Declarations

### Ethical Approval

This study was approved by the University College London Research Ethics Committee (ref: 8227/004). All participants gave written informed consent to take part in the study.

### Data Sharing

Due to the personal and in-depth qualitative accounts given by participants in this study, data has not been made publicly available.

### Author Contributions

All authors contributed to the study conception and design. TM and TJ conducted interviews and analysis under the supervision of JB. The first draft of the manuscript was written by TM and all authors commented on previous versions of the manuscript. All authors read and approved the final manuscript.

#### Acknowledgements

The authors would like to thank the members of the student consultation group who provided additional guidance and feedback throughout this study.

